# Engaging adolescent girls and young women in HIV prevention: A retrospective, observational outcome evaluation of the Eyakho Mo’ghel digital rewards programme in South Africa

**DOI:** 10.64898/2026.07.16.26358227

**Authors:** Alexandra Spyrelis, Cleopatra Sokhela, Admire Chikandiwa, Paul Potsane, Ntombifikile Mtshali

## Abstract

**Introduction:** Adolescent girls and young women (AGYW) in South Africa face disproportionately high HIV incidence, yet uptake and retention in prevention services remain suboptimal. Behavioural economics approaches, including incentive-based models, have shown promise in improving health-seeking behaviours among this population. This study evaluated the Eyakho Mo’ghel (EM) programme, a membership-based digital rewards initiative implemented by Shout-It-Now within the DREAMS HIV prevention framework, to assess its impact on HIV prevention and sexual and reproductive health (SRH) service engagement among AGYW.

**Methods:** A retrospective, observational outcome evaluation was conducted across the full programme implementation period (December 2021–February 2025) in five districts in Gauteng and North West provinces, South Africa. Deidentified clinical and app records for 4,684 EM members were analysed alongside a 1:1 matched comparison group of 4,684 non-members drawn from approximately one million records using stratified random sampling. Outcomes included HIV testing, pre-exposure prophylaxis (PrEP) uptake and persistence, contraceptive use, gender-based violence (GBV) disclosure, and key health indicators. Multivariable logistic and Poisson regression models, adjusted for age and district, were used to examine associations between EM membership, app usage patterns, and outcomes.

**Results:** EM members were over three times more likely to have tested for HIV (OR = 3.16, 95% CI: 2.83–3.54) and tested significantly more frequently than non-members. PrEP initiation was also markedly higher among EM members (OR = 3.15, 95% CI: 2.85–3.48), and persistence beyond the first dispensation was approximately 67% more likely (OR = 1.67, 95% CI: 1.63–1.72). Contraceptive uptake was 75% more likely (OR = 1.75, 95% CI: 1.53–2.01), and EM members were 54% more likely to disclose GBV experiences (OR = 1.54, 95% CI: 1.24–1.91). Sustained app engagement and cumulative point accumulation were consistently associated with improved outcomes. No significant differences in HIV seroconversion, TB screening, or incident pregnancy were observed.

**Conclusions:** A non-monetary, digitally integrated rewards programme was associated with meaningful improvements in HIV prevention service uptake, PrEP persistence, contraceptive use, and GBV disclosure among AGYW. These findings support the integration of incentive-based digital engagement models within combination HIV prevention frameworks, particularly in resource-constrained settings.

## Introduction

Adolescent girls and young women (AGYW) in sub-Saharan Africa, and particularly in South Africa, continue to experience one of the highest HIV incidence rates, underscoring their disproportionate vulnerability [1]. Despite significant advances in biomedical HIV prevention technologies, including pre-exposure prophylaxis (PrEP), uptake and persistence on PrEP, as well as broader retention in HIV prevention services, remain suboptimal among this population [2]. Challenges extend beyond mere access to services, encompassing complex behavioural, structural, and social determinants that influence health-seeking behaviours and sustained engagement with prevention programs [3–5].

Behavioural economics principles have increasingly been applied to HIV prevention, most notably through conditional and unconditional cash transfers, non-monetary rewards, and lottery-based incentives [6]. These approaches leverage the idea that immediate and tangible benefits can counteract present-biased decision-making, thereby encouraging individuals, and particularly adolescents and young people, to initiate and sustain protective behaviours that yield long-term health gains [6]. A growing body of evidence demonstrates that incentive-based interventions are effective in promoting HIV prevention and care outcomes [7]. Conditional and unconditional cash transfers have consistently been shown to reduce HIV risk behaviours and improve service uptake among adolescents and young people, particularly in sub-Saharan Africa [8–10]. Non-monetary rewards and lottery-based incentives have also proven effective in sustaining engagement and motivating continued participation in HIV prevention programmes [6]. Incentive models have further been applied to strengthen linkage to care, adherence to antiretroviral therapy (ART), and uptake of PrEP, with randomized trials demonstrating measurable improvements in retention and health outcomes [7].

Recent evidence suggests that traditional clinic-based approaches to HIV prevention may be insufficient to address the multifaceted barriers faced by AGYW, which include stigma, fear of disclosure, transportation costs, and competing life priorities, among others [8, 9]. Furthermore, the intersection of HIV risk with other health and social vulnerabilities, including unintended pregnancy, sexually transmitted infections (STIs), tuberculosis (TB), and gender-based violence (GBV) necessitates integrated, comprehensive prevention strategies that address the holistic needs of AGYW [1]. The DREAMS (Determined, Resilient, Empowered, AIDS-free, Mentored, and Safe) partnership, implemented across multiple sub-Saharan African countries, exemplified this integrated approach and combined prevention approaches that include biomedical, behavioural, and structural interventions [5, 11].

This paper presents findings from an outcome evaluation of a novel membership-based rewards programme designed to increase AGYW engagement and retention in targeted HIV prevention and sexual and reproductive health (SRH) services in South Africa, called *Eyakho Mo’ghel.* The *Eyakho Mo’ghel programme* was co-developed and delivered by non-governmental organisation, Shout-It-Now, as part of the DREAMS HIV prevention initiative in South Africa.

## Methods

### Setting

Shout-It-Now (hereafter ‘Shout’) delivers a comprehensive suite of innovative, youth-friendly SRH services in high-burden HIV districts in South Africa through the ‘Shout Model’. By integrating peer provider-supported mobile clinic services with a digital ecosystem (including a tailored app, a dedicated call centre, and interactive conversational chatbots), the Shout Model provides a responsive and streamlined service package that ensures seamless access to biomedical, behavioural, psychosocial support and health referrals for AGYW. Grounded in the Theory of Planned Behaviour and the Social Ecological Model [12, 13], Shout emphasises youth engagement and participatory approaches to ensure that young people are actively involved in the design, delivery, and evaluation of services.

As a PEPFAR DREAMS implementing partner, Shout delivered the HIV prevention programme in five districts across the Gauteng and North West provinces between 2019-2025, providing a comprehensive combination prevention package consisting of structured life skills programmes tailored by age group, alongside biomedical services via mobile clinics including HIV testing, PrEP, post-exposure prophylaxis (PEP), linkage to antiretroviral therapy (ART), post-violence care for GBV, screening for STIs and TB, and mental health screening. Demand creation and behaviour change strategies included multi-platform communication involving peer ambassadors, the *Connect Hub* call centre, a mobile app, and social media campaigns.

### Intervention overview

Shout developed the *Eyakho Mo’ghel programme* (EM) as an innovative affinity-based rewards initiative, combining in-person and digital channels with the aim to foster trust, ensure non-judgmental service provision, and strengthen engagement with HIV prevention and SRH services. Conceived as a client-centric membership program, EM was designed through a participatory process that engaged young women directly, analysed clinical management system data, and drew on best practices in youth-friendly programming. Officially launched on World AIDS Day in December 2021 and implemented until February 2025, EM sought to create a safe, supportive community for young women aged 15–24 years, encouraging and celebrating healthy behaviours while providing access to resources, information, and peer support. By embedding rewards within service uptake, the programme aimed to increase retention in prevention services and build a sense of belonging among participants, thereby enhancing their willingness to seek support, restart services, or refer peers.

Central to the intervention was the integration of the *iSHOUT!* app, a data-free digital platform offering members health information, a mobile clinic locator, appointment reminders, economic strengthening resources, and virtual chat functions. Eligibility for EM required participation in at least one life skills session and receipt of a biomedical HIV or SRH service under DREAMS, after which clients were invited to enrol through digital or mobile clinic-based channels. Upon joining, members received symbolic and practical items, including an EM scarf and a reusable menstrual cup, alongside access to the *iSHOUT!* app and the opportunity to earn points for healthy behaviours such as HIV testing, PrEP initiation and continuation, contraceptive use, participating in life skills programmes, and survey completion. Points could be redeemed in a Marketplace for cosmetics, electronics, or retail vouchers. The programme’s design incorporated a structured exit process at age 25, ensuring sustainability and turnover while maintaining access to informational resources. Through this multilayered approach, EM sought not only to increase uptake and retention in HIV prevention and SRH services but also to contribute to broader outcomes such as improved health literacy, economic empowerment, and ultimately, reduced HIV transmission among young women.

### Study design

The study employed a retrospective, observational outcome evaluation of the EM programme across its full implementation period (December 2021-February 2025). The evaluation focused on two primary objectives, namely: i) assessing the impact of the EM membership-based rewards programme on uptake of and retention in SRH services among AGYW; and ii) examining the impact of the EM programme on health outcomes among AGYW. Secondary data sources were utilised, including identified clinic records captured in Shout’s clinical management system throughout the implementation period, and usage data from the *iSHOUT!* mobile application. These datasets provided information on service utilisation, engagement patterns, and programme outcomes, enabling analysis of both behavioural and clinical indicators.

### Study population

Secondary analysis of deidentified data was conducted to assess key outcomes. Deidentified records for 4,684 EM members were extracted from Shout’s digital clinical management system and the *iSHOUT!* mobile application, for the full implementation period (1 December 2021 to 28 February 2025). These records were merged into a unified database of behavioural and clinical outcomes using a unique identifier.

A comparison group of 4,684 non-EM members (i.e. a 1:1 match) was randomly selected from the clinical management system for the same period. Stratified random sampling was applied to the full clinical management system database of approximately 1,000,000 records. To maintain reproducibility, the random number generator was initialized using a fixed seed in STATA (set seed 12345). The sampling fraction was calculated as 4,684 ÷ 1,000,000 = 0.0046 (0.4684%). Each stratum’s sample size was determined proportionally to its population size. Random selection was performed in STATA using the “*sample”* command with the *“by ()”* option to select 0.4684% of records from each stratum. This ensured that the final sample preserved the overall distribution of the stratification variable. The final dataset was verified to confirm the sample size and proportional distribution across strata.

### Data analysis

Descriptive statistics summarized baseline characteristics: categorical variables as frequencies and percentages, and continuous variables as means with standard deviations (SD) or medians with interquartile ranges (IQR), depending on distribution. Group comparisons employed chi-square or Fisher’s exact tests for categorical variables, and independent-samples t-tests or Wilcoxon rank-sum tests for continuous variables, as appropriate. Associations between exposures and outcomes were examined using regression models: logistic regression for binary outcomes, and Poisson or negative binomial regression for count outcomes, depending on distributional properties and overdispersion. Multivariable models were adjusted for age and district, identified a priori as confounders. Analyses of programme outcomes were conducted using Stata version 18.1 (StataCorp LLC, College Station, TX, USA). Statistical significance was defined as p<0.05 (two-sided).

### Ethics

This study formed part of a broader evaluation of the DREAMS programme implemented by Shout, which was approved by the South African Medical Association Research Ethics Committee (SAMAREC) (280808016/073/2025). It involved a secondary analysis of deidentified clinic service and *iSHOUT!* app records. Participants provided informed consent for both clinical enrolment and onboarding onto *iSHOUT!*, permitting data use for research and programme improvement.

## Results

### Participant characteristics

The study population comprised 4,684 EM members matched to 4,684 comparison participants. Significant differences were observed in age distribution, with a higher proportion of EM members aged 15–19 years (40.9% vs. 23.4%) and fewer aged 20–24 years (59.1% vs. 76.6%; *p* < 0.001). Provincial distribution also differed slightly between groups, though both were predominantly from Gauteng (72.5% vs. 72.2%; *p* < 0.001). HIV status was similar across groups, with the majority testing negative (97.5% EM vs. 97.7% comparison; *p* = 0.126). Pregnancy status did not differ significantly between groups (14.6% vs. 14.3%; *p* = 0.964) (Table 1).

**Table 1.**
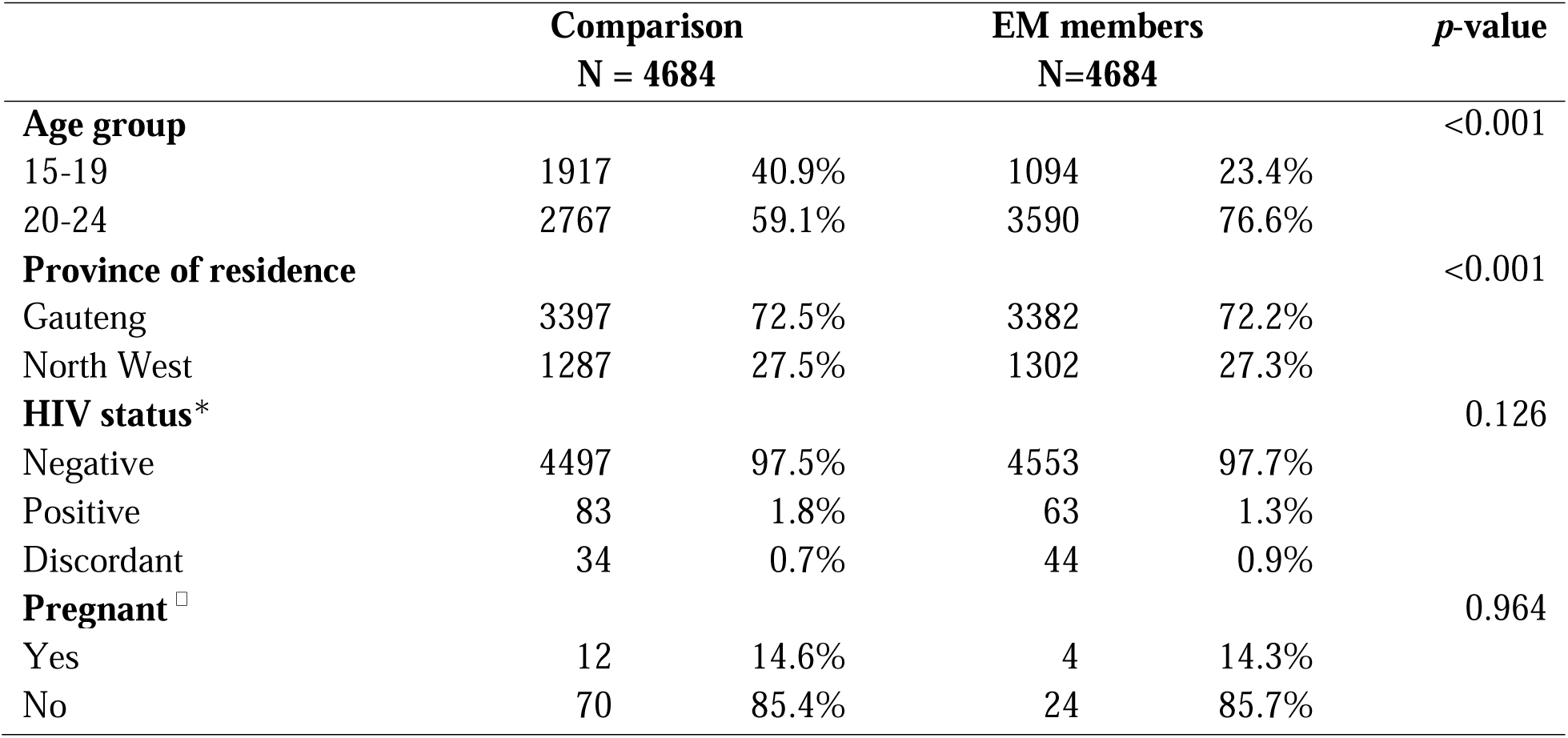
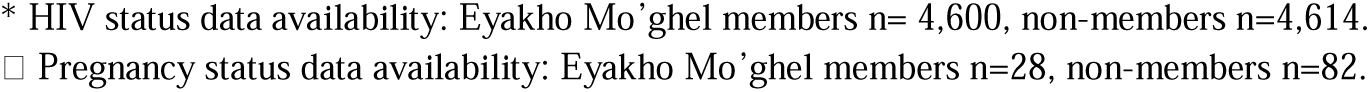
Demographic characteristics of study population.

|  | Comparison<br>N = 4684 |  | EM members<br>N=4684 |  | p-value |
| --- | --- | --- | --- | --- | --- |
| **Age group** |  |  |  |  | <0.001 |
| 15-19 | 1917 | 40.9% | 1094 | 23.4% |  |
| 20-24 | 2767 | 59.1% | 3590 | 76.6% |  |
| **Province of residence** |  |  |  |  | <0.001 |
| Gauteng | 3397 | 72.5% | 3382 | 72.2% |  |
| North West | 1287 | 27.5% | 1302 | 27.3% |  |
| **HIV status\*** |  |  |  |  | 0.126 |
| Negative | 4497 | 97.5% | 4553 | 97.7% |  |
| Positive | 83 | 1.8% | 63 | 1.3% |  |
| Discordant | 34 | 0.7% | 44 | 0.9% |  |
| **Pregnant<sup>□</sup>** |  |  |  |  | 0.964 |
| Yes | 12 | 14.6% | 4 | 14.3% |  |
| No | 70 | 85.4% | 24 | 85.7% |  |

\* HIV status data availability: Eyakho Mo'ghel members n= 4,600, non-members n=4,614.
□ Pregnancy status data availability: Eyakho Mo'ghel members n=28, non-members n=82.

### Uptake of biomedical services

Table 2 presents uptake of SRH services by study group. While the uptake of HIV testing was almost universal (99% among both groups), EM members tested significantly more frequently than non-members (mean number of tests of 2.7 and 1.6 respectively, *p* < 0.001). EM members were more than three times more likely to have taken an HIV test compared to non-members (OR = 3.16, 95% CI: 2.83– 3.54).

**Table 2.**
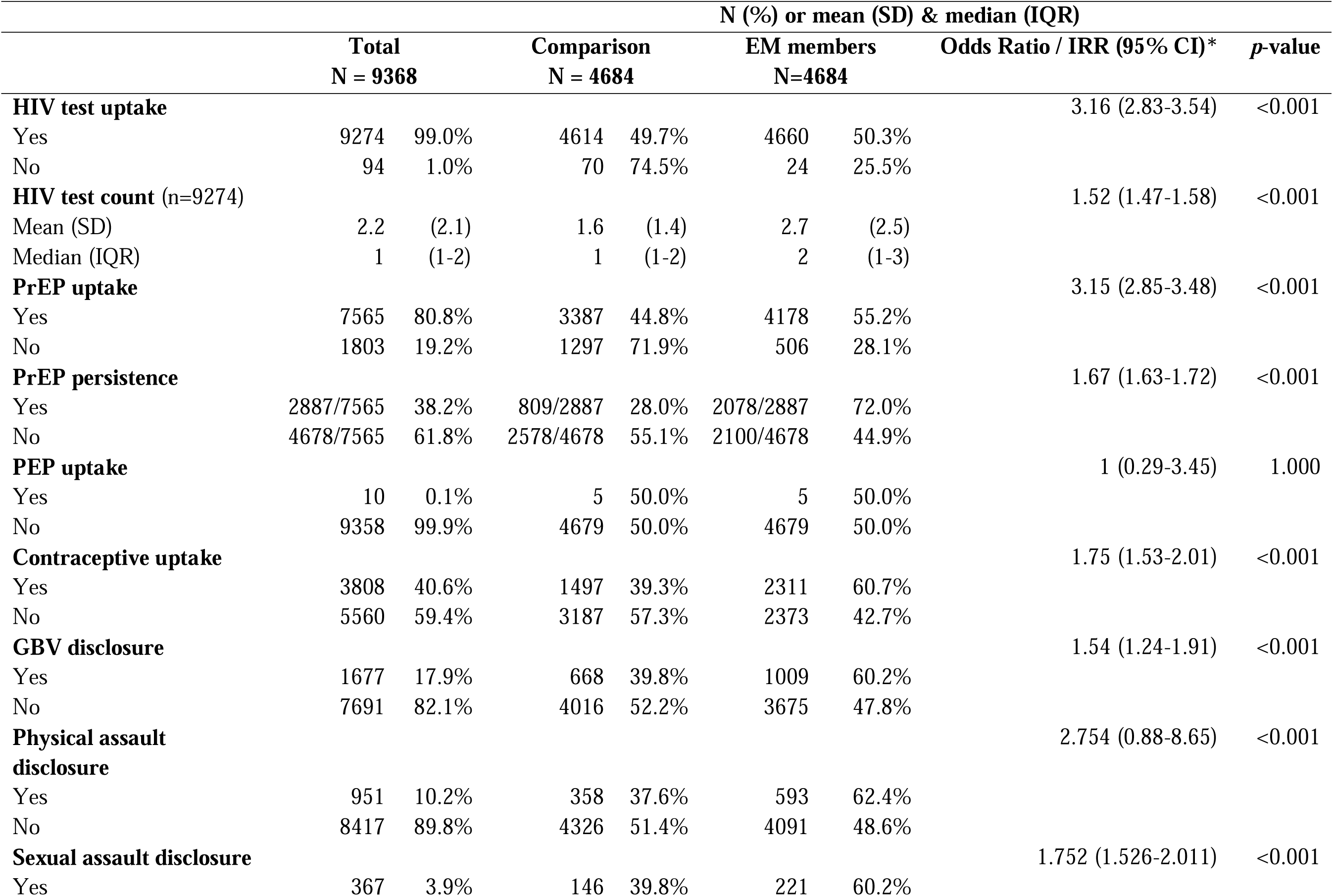
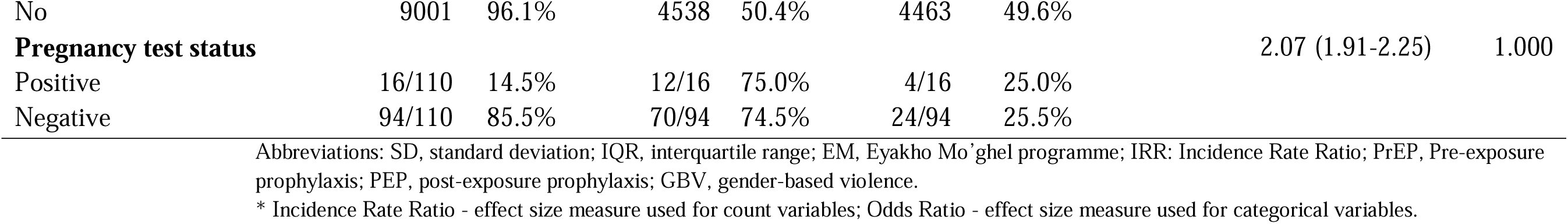
Uptake of SRH services among study groups, 1 December 2021-28 February 2025.

PrEP uptake was also significantly higher among EM members than non-members (55.2% and 44.8% respectively, *p* < 0.001). EM members were over three times more likely to initiate PrEP (OR = 3.15, 95% CI: 2.85–3.48). Further, a significantly higher proportion of EM members (72%) persisted on PrEP beyond the first issue versus non-members (28%) (*p* < 0.001). EM members were about 67% more likely to persist with PrEP (OR = 1.67, 95% CI: 1.63–1.72). PEP use was low among both groups, with no significant difference observed (*p* = 1.000).

Furthermore, contraceptive uptake was greater among EM members (60.7%) than non-members (39.3%) (*p* < 0.001). EM members were about 75% more likely to use contraceptives (OR = 1.75, 95% CI: 1.53– 2.01). Disclosures of GBV were higher among EM members (60.2%) than non-members (39.8%) (*p* < 0.001), with EM members about 54% more likely to disclose GBV (OR = 1.54, 95% CI: 1.24–1.91). Identified cases were predominantly for physical assault (62.4% of EM members) and sexual assault (60.2% of EM members). The number of pregnancy tests conducted was low among both groups, with no difference in positivity rates (*p* = 1.000).

### Impact on health outcomes

The STI screening was marginally higher (62%) among EM members compared to non-EM members (38%, *p* = 0.064). While some differences were observed between EM members and non-members on the other key health outcomes, none of them were statistically significant (Table 3).

**Table 3.**
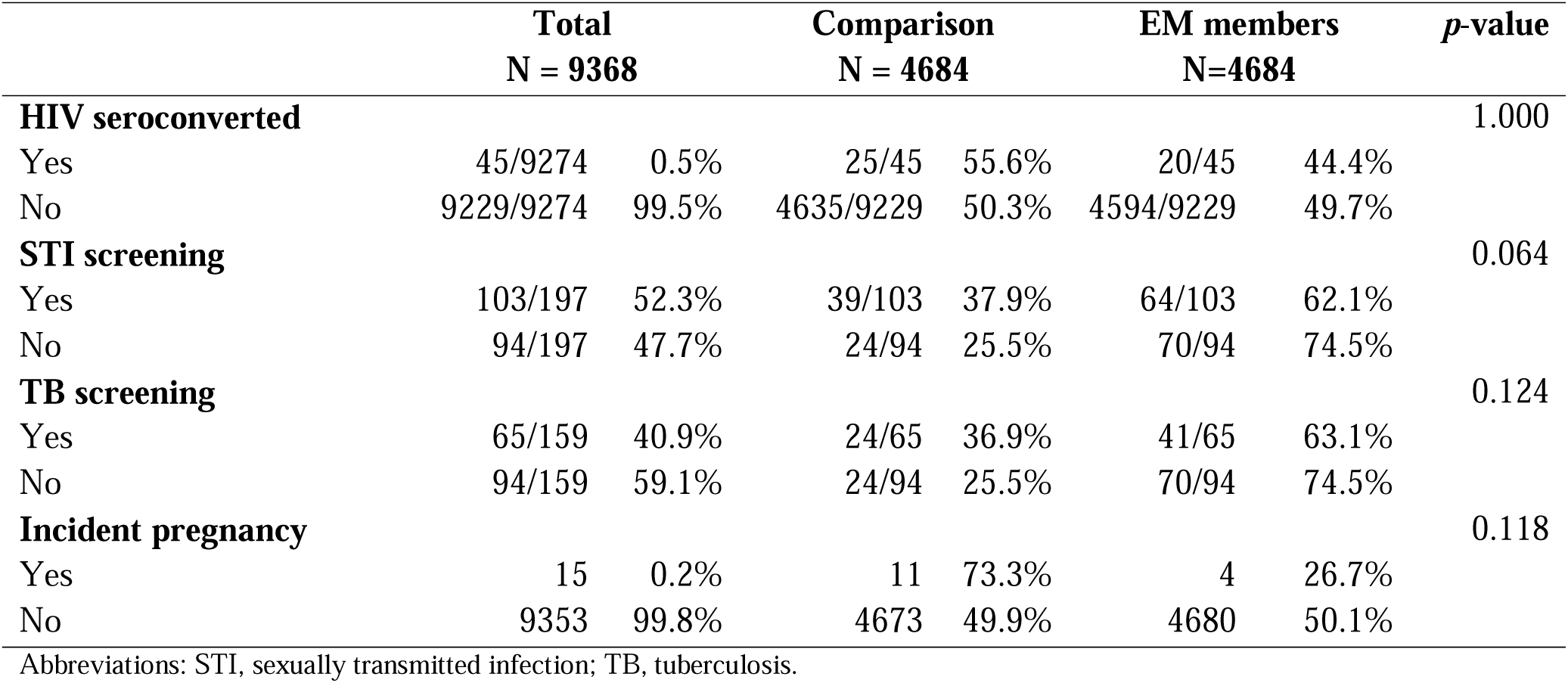
Health outcomes among study groups, December 2021-February 2025.

|  | Total<br>N = 9368 |  | Comparison<br>N = 4684 |  | EM members<br>N=4684 |  | p-value |
| --- | --- | --- | --- | --- | --- | --- | --- |
| **HIV seroconverted** |  |  |  |  |  |  | 1.000 |
| Yes | 45/9274 | 0.5% | 25/45 | 55.6% | 20/45 | 44.4% |  |
| No | 9229/9274 | 99.5% | 4635/9229 | 50.3% | 4594/9229 | 49.7% |  |
| **STI screening** |  |  |  |  |  |  | 0.064 |
| Yes | 103/197 | 52.3% | 39/103 | 37.9% | 64/103 | 62.1% |  |
| No | 94/197 | 47.7% | 24/94 | 25.5% | 70/94 | 74.5% |  |
| **TB screening** |  |  |  |  |  |  | 0.124 |
| Yes | 65/159 | 40.9% | 24/65 | 36.9% | 41/65 | 63.1% |  |
| No | 94/159 | 59.1% | 24/94 | 25.5% | 70/94 | 74.5% |  |
| **Incident pregnancy** |  |  |  |  |  |  | 0.118 |
| Yes | 15 | 0.2% | 11 | 73.3% | 4 | 26.7% |  |
| No | 9353 | 99.8% | 4673 | 49.9% | 4680 | 50.1% |  |

Abbreviations: STI, sexually transmitted infection; TB, tuberculosis.

### Impact of app usage patterns

An overview of EM members’ usage patterns of the *iSHOUT!* app is provided in Table 4. EM participant app users were 21.1 years old (*SD* = 2.1) on average and almost all were active during the programme period (99%). EM members spent an average of 7.4 months (*SD* = 7.8) on the app during this time, earning an average of 118.3 points (*SD* = 64.1) and redeeming an average of 45.1 points (*SD* = 302.8) across a mean of 25.5 transactions (*SD* = 61.3).

**Table 4.** Characteristics of EM members based on backend app data (n=4673)*

| Description | Mean (SD) | Median (IQR) |
| --- | --- | --- |
| Age – years | 21.1 (2.1) | 21 (20-23) |
| Duration on app – months <sup>□</sup> | 7.4 (7.8) | 4.2 (1.3-12.0) |
| Current points available | 85.9 (56.3) | 84.7 (45.3-114.7) |
| Total points earned | 118.3 (64.1) | 104.6 (82.7-143.1) |
| Total points redeemed | 45.1 (302.8) | 0 (0-70) |
| Total allocations | 21.4 (11.4) | 19 (14-27) |
| Total redemptions | 0.9 (1.9) | 0 (0-1) |
| Total transactions | 25.5 (61.3) | 22 (16-31) |

\* Backend app data were available for only 4,673 out of the 4,684 EM members included in this analysis.
<sup>□</sup> Duration on app data were complete for 4,672 members.

Patterns of app usage were significantly associated with HIV prevention outcomes among EM members. With regard to HIV testing, longer duration of app engagement and higher point accumulation were consistently linked to increased testing frequency (*p* < 0.001), while frequent transaction episodes (defined as the number of times points were awarded, regardless of the amount) were negatively associated with testing (*p* < 0.001) (Table 5).

**Table 5.** Impact of app usage patterns on HIV testing count.

| Description | B (95% CI) | p-value |
| --- | --- | --- |
| Duration in months | 0.13 (0.12-1.14) | <0.001 |
| Total points earned | 0.01 (0.008-1.02) | <0.001 |
| Total points redeemed | 0.005 (0.005-0.008) | 0.03 |
| Current points | -0.02 (-0.01-0.02) | 0.33 |
| Total allocations | -0.0006 (-0.02-0.22) | 0.99 |
| Total redemptions | 0.05 (-0.003-1.02) | 0.07 |
| Total transactions | -0.02 (-0.04 to -0.03) | 0.02 |
| Age | 0.09 (0.07-0.12) | <0.001 |

Note: Coefficients reflect change in HIV testing count per unit increase in predictor. Models adjusted for age and district.

For PrEP uptake, app usage patterns showed mixed associations (Table 6). Each point earned on the app was associated with a 3% increase in the odds of PrEP initiation (OR = 1.03, 95% CI: 1.02–1.04, *p* < 0.001), while each point allocation episode was linked to a 9% reduction in uptake (OR = 0.91, 95% CI: 0.87-0.95, *p* < 0.001). Other usage variables, including duration of app engagement, current points balance, total points redeemed, redemption episodes, and transaction episodes, did not show significant associations.

**Table 6.** Impact of app usage patterns on PrEP uptake.

| Description | Odds Ratio (95% CI) | p-value |
| --- | --- | --- |
| Duration in months | 0.99 (0.98-1.01) | 0.77 |
| Total points earned | 1.03 (1.02-1.04) | <0.001 |
| Total points redeemed | 0.99 (0.98-1.01) | 0.98 |
| Current points | 1.00 (0.99-1.01) | 0.91 |
| Total allocations | 0.91 (0.87-0.95) | <0.001 |
| Total redemptions | 0.99 (0.84-1.18) | 0.96 |
| Total transactions | 1.02 (0.98-1.07) | 0.21 |
| Age | 1.03 (0.98-1.08) | 0.22 |
| District | 1.11 (1.05-1.17) | <0.001 |

Note: Odds ratios reflect change in likelihood of PrEP uptake per unit increase in predictor. Models adjusted for age and district.

Persistence in PrEP use was positively associated with sustained app engagement and point accumulation (Table 7). Each additional month of app usage increased the odds of persistence by 9% (OR = 1.09, 95% CI: 1.08–1.10, *p* < 0.001), while each point earned increased persistence by 2% (OR = 1.02, 95% CI: 1.01–1.03, *p* < 0.001) and each point redeemed by 1% (OR = 1.01, 95% CI: 1.04–1.02, *p* < 0.001). Current points balance also showed a modest positive association. In contrast, point allocation episodes were linked to a 4% reduction in persistence (OR = 0.96, 95% CI: 0.92–0.99, *p* < 0.001). Other usage variables, including redemption episodes and transaction activity, were not significantly associated.

**Table 7.** Impact of app usage patterns on PrEP persistence.

| Description | Odds Ratio (95% CI) | *p*-value |
| --- | --- | --- |
| Duration in months | 1.09 (1.08-1.10) | <0.001 |
| Total points earned | 1.02 (1.01-1.03) | <0.001 |
| Total points redeemed | 1.01 (1.04-1.02) | <0.001 |
| Current points | 1.01 (1.005-1.01) | 0.02 |
| Total allocations | 0.96 (0.92-0.99) | 0.01 |
| Total redemptions | 1.04 (0.93-1.15) | 0.51 |
| Total transactions | 0.97 (0.94-1.00) | 0.10 |
| Age | 1.11 (1.07-1.15) | <0.001 |

Note: Odds ratios reflect change in likelihood of PrEP persistence per unit increase in predictor. Models adjusted for age and district.

Additional analyses explored the impact of app usage patterns on contraceptive service uptake and disclosure of violence. These findings are presented in Tables 8-11 in Appendix 1. In summary, longer app engagement and point accumulation were positively associated with contraceptive service uptake, with each additional month of usage increasing the odds by 4% (OR = 1.04, 95% CI: 1.02–1.05, *p* < 0.001) and each point earned by 1% (OR = 1.01, 95% CI: 1.004–1.02, *p* < 0.001) (Table 8). For GBV disclosure, only duration of app use showed a significant association, with each month linked to a 2% increase in disclosure (OR = 1.02, 95% CI: 1.01–1.03, *p* < 0.001) (Table 9). Physical assault disclosure was positively associated with longer app engagement (OR = 1.02, 95% CI: 1.01–1.03, *p* < 0.001) and point allocation episodes (OR = 1.04, 95% CI: 1.01–1.07, *p* = 0.03) (Table 10). Finally, sexual assault disclosure was positively associated with points earned (OR = 1.01, 95% CI: 1.01–1. 02, *p* = 0.006) and transaction episodes (OR = 1.04, 95% CI: 1.01–1. 08, *p* = 0.03), but negatively associated with points redeemed (OR = 0.99, 95% CI: 0.98–0.99, *p* = 0.03) and current point balance (OR = 0.99, 95% CI: 0.98–0.99, *p* = 0.009) (Table 11).

## Discussion

Our evaluation of the Eyakho Mo’ghel programme, implemented by Shout-It-Now between 2021–2025 within the broader DREAMS HIV prevention programme, demonstrates that a rewards-based, technology-driven approach meaningfully improved AGYW uptake and retention in HIV prevention and SRH services, while strengthening GBV risk identification and response.

EM members were more than three times more likely to get tested for HIV and tested significantly more frequently than non-members. Achieving high coverage of repeat HIV testing among at-risk individuals remains a persistent challenge in the prevention cascade in sub-Saharan Africa and is critical for achieving the UNAIDS 95-95-95 targets [14]. Our findings align with the HPTN 068 trial, which showed that conditional cash transfers increased HIV testing among young women in South Africa [15], but extend this evidence by demonstrating comparable effects through a non-monetary digital model. Lee et al.’s [16] systematic review of incentives for HIV and STI testing found that non-monetary incentives were equally effective as monetary ones, and specifically that incentives offered in non-clinical settings were more impactful; a feature of the EM model that may have contributed to the effects observed.

EM members were also more than three times more likely to initiate PrEP and approximately 67% more likely to persist on it than young women in the comparison group. These findings are particularly striking given the steep attrition in PrEP use consistently documented in real-world implementation settings [17, 18]. Celum et al.’s [19] trial of financial incentives tied to tenofovir levels among young women in South Africa found only a non-significant trend toward improved adherence at month three, which attenuated by month twelve following incentive withdrawal and reduced visit frequency, pointing to the importance of sustained structured engagement alongside any incentive. Our findings suggest that the stronger persistence effect observed in EM may reflect the additive contribution of sustained programme contact, peer community, and progressive reward accumulation. This is consistent with Chen-Charles et al.’s [18] scoping review of PrEP uptake among AGYW in sub-Saharan Africa, which identified social support, regular provider contact, and differentiated service delivery as the most consistent facilitators of long-term persistence, which were all structurally embedded in the EM model.

EM membership was also associated with a 75% increase in the likelihood of contraceptive uptake. Given well-documented demand-side barriers including partner disapproval, side effect misconceptions, and sociocultural fertility norms [20], the EM programme’s trusted relationships, digital engagement, and non-monetary rewards may have created enabling conditions that access-focused interventions alone cannot. While the broader cash transfer literature on contraceptive outcomes remains mixed [21], the consistent finding that incentive-based programmes improve reproductive health service engagement when embedded within social support structures, is relevant to interpreting the contraceptive uptake gains observed in EM.

EM members were 54% more likely to disclose experiences of GBV, primarily physical and sexual assault, than non-members. This is particularly important given the well-established bidirectional relationship between GBV and HIV risk, and the relatively low rates of GBV identification within HIV prevention programmes [22, 23]. Higher disclosure rates among EM members likely reflect the trust cultivated through sustained engagement, recognition by programme navigators, and the non-stigmatising framing of the programme - conditions that are difficult to achieve through single-contact clinical interactions.

Despite these promising findings, no measurable reduction in HIV seroconversions was observed. This is not unexpected: a four-year programme period is likely insufficient to detect incidence-level change, particularly in a comparison group that also received DREAMS services. The lack of significant differences in TB and pregnancy rates may partly be explained by the fact that EM members were more likely to seek care and get tested, meaning more cases were detected among them simply because they attended services more often, not necessarily because they were more unwell. This makes it difficult to interpret these comparisons as true differences in disease rates. More broadly, improvements in individual service uptake do not automatically translate into fewer HIV infections at the population level. This is because HIV transmission also depends on the behaviour and HIV status of sexual partners, who are largely outside the reach of a programme focused on AGYW alone. UNAIDS [24] has identified reaching the male sexual partners of AGYW with HIV testing, treatment, and prevention services as a critical and underutilised strategy for reducing HIV incidence among young women, underscoring the need to embed rewards-based AGYW engagement models within broader combination prevention frameworks that extend to partners and communities.

Our findings resonate with Zhang et al.’s [7] global review, which concluded that monetary incentives often show stronger short-term effects while non-monetary incentives may be more sustainable and less prone to unintended consequences. Our results support and refine this distinction: earning and redeeming points were consistently associated with positive outcomes, while transactional activity and point allocation episodes showed mixed or negative associations. This suggests that incentive design matters as much as incentive type, and that poorly structured rewards can inadvertently dampen motivation.

This study has several limitations. The observational design raises the possibility of residual confounding. The relatively short follow-up period limited the ability to detect HIV incidence-level differences. Small sample sizes for TB and STI outcomes power to detect any significant differences. Since EM was implemented concurrently with other DREAMS campaigns, exclusive attribution of observed effects to the EM intervention is not possible, though this real-world context offers practical lessons for policymakers on how incentive-based models can add value within multi-layered prevention frameworks. Categorical age measurement constrained demographic precision, and questions remain about digital equity, particularly for the most marginalised AGYW with limited smartphone ownership, data access, or digital literacy.

## Conclusions

Sustained and active engagement with a non-monetary, technology-driven rewards programme was positively associated with improved HIV testing, PrEP initiation and persistence, contraceptive uptake, and GBV disclosure among AGYW. This evaluation provides novel, policy-relevant evidence that complements existing trials, demonstrating that the architecture of the incentive, the quality of programme relationships, and the sustainability of digital engagement may matter as much as the monetary value of the reward itself. As the field moves toward implementation at scale in the context of tightening donor funding, this evidence supports the case for non-monetary, digitally integrated incentive models as a feasible and potentially cost-effective component of combination HIV prevention for AGYW.

## Authors’ contributions

CS, AC, PP, NM, and AS conceptualised the study and study design. AS and CS were responsible for the data handling. AC led the statistical analysis with input from AS and CS. AS led the writing of the manuscript with input from CS, AC, PP, and NM. All authors had access to all study data and had final responsibility for the decision to submit for publication.

## Data Availability

De-identified individual participant data and the accompanying data dictionary will be made available upon reasonable written request to the Principal Investigator, subject to approval of a formal proposal and completion of a signed Data Access Agreement. Requests must specify the intended research purpose and planned analyses. Additional study documents, including the study protocol and statistical analysis plan, may be shared where appropriate. Data will be available from the time of publication and for a period of five years thereafter. Data will be shared via secure institutional channels and without investigator support unless otherwise agreed.

## Acknowledgements

We are deeply grateful to all members of the Eyakho Mo’ghel programme team at Shout-It-Now for their invaluable contributions. In particular, we acknowledge the virtual engagement team, led by Pierrette Kengela, who managed the programme and provided critical insights for the evaluation, together with Wesley Ramaselele, Thandeka Thwala, Vinoria Jacobs, and Palesa Lehoko. We thank Ranganai Makiwa and Yonwaba Pezisa for their role in conceptualising the work; Vuyi Skiti and team for retrieving data from Shout’s clinical management system; and Neetin Daya for contributing to study insights. We also extend our appreciation to the broader implementation team whose dedication ensured the programme’s success, including the peer ambassadors, clinicians at the mobile clinics, and staff at the Connect Hub call centre.

## List of abbreviations

AGYW: Adolescent girls and young women
ART: Antiretroviral therapy
DREAMS: Determined, Resilient, Empowered, AIDS-free, Mentored, and Safe
EM: Eyakho Mo’ghel programme
GBV: Gender-based violence
IQR: Interquartile range
NGO: Non-governmental organisation
PEP: Post-exposure prophylaxis
PEPFAR: President’s Emergency Plan for AIDS Relief
PrEP: Pre-exposure prophylaxis
SD: Standard deviation
SRH: Sexual and reproductive health
STI: Sexually transmitted infection
TB: Tuberculosis

